# A population-specific missense variant rs1597000001 in *CETP* promotes a favorable lipid profile and reduces CETP activity

**DOI:** 10.1101/2021.09.11.21263438

**Authors:** Jaye Moors, Mohanraj Krishnan, Nick Sumpter, Riku Takei, Matt Bixley, Murray Cadzow, Tanya J. Major, Amanda Phipps-Green, Ruth Topless, Marilyn Merriman, Malcolm Rutledge, Ben Morgan, Jenna C. Carlson, Jerry Z. Zhang, Emily M. Russell, Guangyun Sun, Hong Cheng, Daniel E. Weeks, Take Naseri, Muagututi‘a Sefuiva Reupena, Satupa‘itea Viali, John Tuitele, Nicola L. Hawley, Ranjan Deka, Stephen T. McGarvey, Janak de Zoysa, Rinki Murphy, Nicola Dalbeth, Lisa Stamp, Mele Taumoepeau, Frances King, Philip Wilcox, Sally McCormick, Ryan L. Minster, Tony R. Merriman, Megan Leask

## Abstract

Sequencing of *CETP* in Māori and Pacific peoples identified a common (MAF ∼2.4%-5.4%) population-specific missense variant (rs1597000001, *CETP*:c.530C>T p.Pro177Leu) that associates with higher HDL-C levels (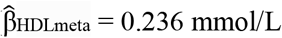 [95% CI 0.211; 0.260]) and lower LDL-C (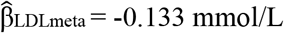 [95% CI -0.209; -0.058]). In a subsample of the study cohort (n = 11), heterozygous carriers of the population-specific variant had lower plasma CETP activity (*P* = 0.028). Our study identifies a population-specific missense variant in *CETP* which lowers CETP activity with an effect on HDL-C that is comparable to Mendelian *CETP* loss-of-function mutations.

---

Dyslipidaemia, defined as elevated total or low-density lipoprotein cholesterol (LDL-C) levels, triglycerides, and/or lower levels of high-density lipoprotein cholesterol (HDL-C), is an established risk factor for metabolic diseases such as cardiovascular disease, type 2 diabetes and gout^1^. In gout, elevated triglycerides may contribute to increased urate levels ^2^, and elevated very low-density-lipoprotein (VLDL) associates with gout in Aotearoa New Zealand (NZ) Māori and Pacific peoples (herein defined as peoples of Micronesian, Melanesian and Polynesian ethnicity) ^3^. Dyslipidaemia is more prevalent in Māori and Pacific populations ^4,5^, and a higher proportion of Pacific nation ancestry has been associated with lower HDL-C ^6,7^. Consistent with heritability estimates of HDL-C levels (40%–60%) ^8,9^, the prevalence of dyslipidaemia in Māori and Pacific populations suggests that genetics contributes to an atherogenic lipid profile ^10^. While socio-economic inequities contribute to the development and increased impact of metabolic diseases ^10,11^, research on population-specific genetic variants associated with metabolic traits is important to provide insights into the genetic determinants of phenotypic differences between populations, as well as furthering genomic justice ^12^.

One locus with genetic variation that consistently associates with lipid levels in multiple population groups, including Pacific peoples, is at the gene that encodes cholesteryl ester transfer protein: CETP ^13-16^. CETP modifies lipid levels by mediating the bi-directional transfer of cholesterol esters from atheroprotective HDL to atherogenic VLDL in exchange for triglycerides, and vice versa. Various common polymorphisms including the *CETP* promoter variant ‘-629 C/A’ (rs1800775), intronic ‘Taq1B’ variant (rs708272), missense variant p.Ile405Val (rs5882) and loss-of-function variant p.Asp459Gly (rs2303790) have been associated with serum lipid profiles, CETP levels and/or CETP activity, and cardiovascular disease and pharmacological response to CETP inhibitors ^17-20^. In a GWAS for lipids in Samoans (using samples analyzed in greater depth here), the *CETP* locus was identified as the most significantly associated locus for HDL-C levels (rs289708, *P* = 1.19 × 10^−11^) ^15^. Given the previously reported associations with lipid levels and variation at *CETP* in Pacific peoples^15,21^, we used a discovery and replication study design ^22^ to investigate whether population-specific genetic variation in *CETP* could contribute to HDL-C and CETP activity in Māori and Pacific populations.

## RESULTS

### Discovery of a novel *CETP* missense variant rs1597000001 in Māori and Pacific people

We extracted the gene region of *CETP* (∼22 Kb) from whole-genome sequence data for 55 individuals of Māori and Pacific ethnicity, and identified a common missense variant in exon 6 of the *CETP* gene (dbSNP (build 155) rs1597000001, *CETP*:c.530C>T, p.Pro177Leu). The minor allele frequency (MAF) was 3.4% in the 55 discovery genomes and 0.0036% in GnomAD ^23^. A CADD score ^24^ of 24.4 placed the variant in the top 1% of predicted deleteriousness and a GERP score ^25^ of 4.4 indicated that the variant is conserved in mammals (Figure 1A, CADD and GERP tracks, respectively). The p.Pro177Leu amino acid substitution caused by the rs1597000001 T-allele corresponds to amino acid position 160 in the mature protein structure of CETP ^26^ owing to the first 17 amino acid residues of the CETP sequence consisting of the signal peptide ^27^ (Figure 1B).

**Figure 1.**
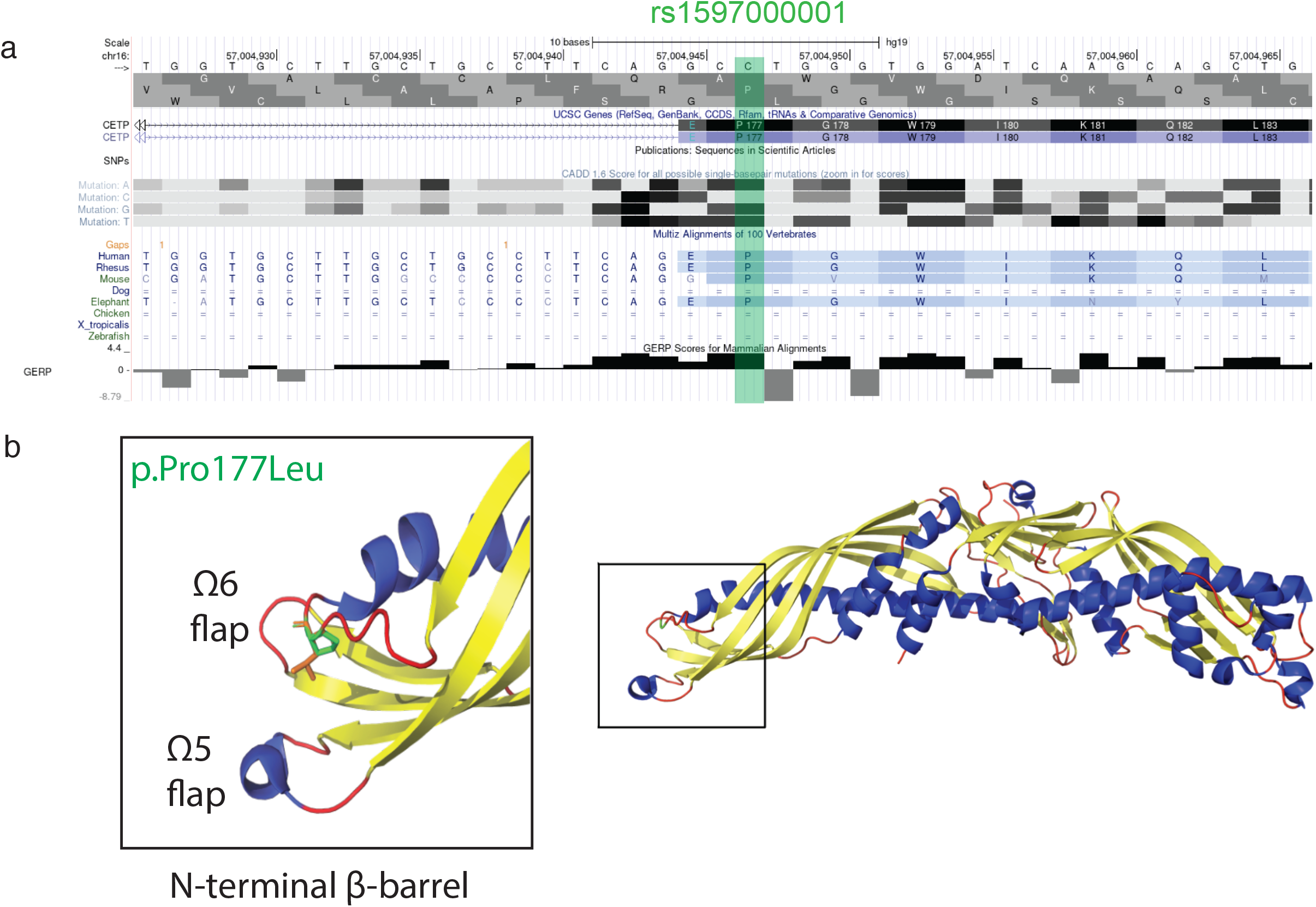
The population-specific rs1597000001 variant changes the amino acid sequence of CETP. A. The CADD, GERP and Multiz UCSC track alignment at rs1597000001 for which the substitution creates a non-synonymous amino acid residue change (p.Pro177Leu). 3D schematic of p.Pro177Leu (P160L) in the mature CETP protein structure in the N-terminal barrel domain (left) of CETP. The CETP protein structure (10.2210/pdb2OBD/pdb) ^26^ was obtained from The Protein Databank (https://www.rcsb.org/) ^52^ and illustrated using PyMOL. CETP, cholesteryl ester transfer protein. CADD, Combined Annotation Dependent Depletion scores. GERP, Genomic Evolutionary Rate Profiling.

### rs1597000001 associates with higher HDL-C

We genotyped rs1597000001 in a discovery cohort of 2,270 Māori and Pacific individuals living in Aotearoa NZ. Demographic and biochemical characteristics of the participants are summarized in Table 1. A similar MAF to the 55 Māori and Pacific genome sequences was observed (MAF = 3.5%), and the MAF ranged from 3.2% - 5.4% in the Pacific nation subgroups (stratified by self-reported Pacific nation of grandparental ethnicity) (Aotearoa NZ NZ Māori MAF 3.4%, Aotearoa NZ Cook Island Māori MAF 3.8%, Aotearoa NZ Samoan MAF 4.0%, Aotearoa NZ Tongan MAF 4.2%, Aotearoa NZ Niuēan MAF 5.4% and Aotearoa NZ Other/Mixed MAF 4.5%). People of Pukapukan ethnicity were monomorphic for the C allele of rs1597000001 (Table 2). The MAF was 2.4% in the Ngāti Porou Hauora group. rs1597000001 did not significantly deviate from Hardy-Weinberg equilibrium (HWE) in any of the Aotearoa NZ Pacific nation subgroups or the Ngāti Porou Hauora group (Table 2: *P* > 0.05, all datasets).

**Table 1.**
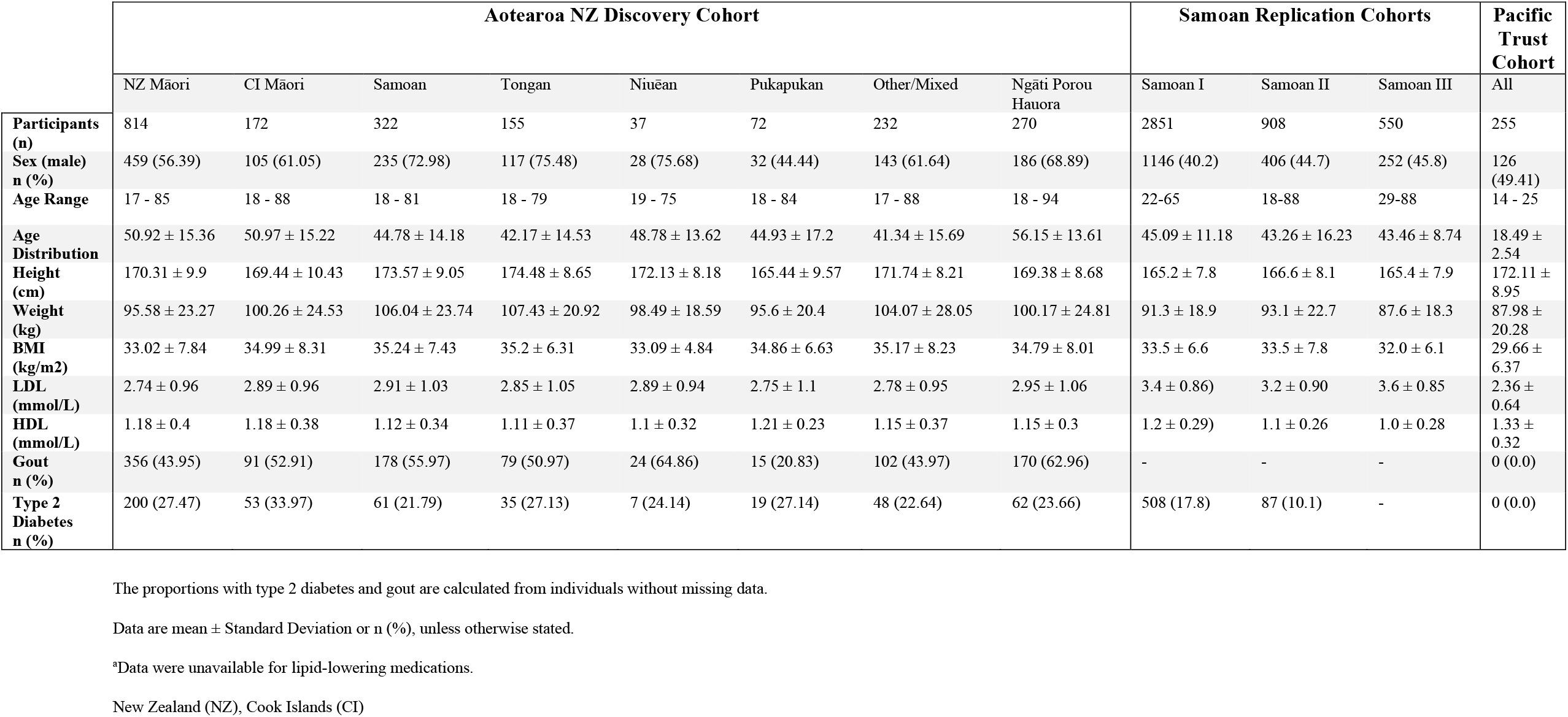
Baseline characteristics of the cohorts.

**Table 2.** Minor Allele Frequency and Hardy-Weinberg Equilibrium of rs1597000001.

|  | Aotearoa NZ<br>Discovery Cohort |  |  |  |  |  |  |  | Samoa<br>Replication Cohorts |  |  | Pacific Trust Otago<br>Cohort |  |
| --- | --- | --- | --- | --- | --- | --- | --- | --- | --- | --- | --- | --- | --- |
|  | NZ<br>Māori | CI<br>Māori | Samoa | Tongan | Niuean | Pukapukan | Other/Mixed | Ngāti<br>Porou<br>Hauora | Samoa I | Samoa II | Samoa III | Polynesian | Non-Polynesian<br>Pacific peoples |
| **Total (n)** | 814 | 172 | 322 | 155 | 37 | 72 | 232 | 270 | 2851 | 908 | 550 | 220 | 35 |
| **CC (n)** | 760 | 159 | 297 | 143 | 33 | 72 | 212 | 257 | 2600 | 825 | 497 | 201 | 35 |
| **(%)** | (93.4) | (92.4) | (92.2) | (92.3) | (89.2) | (100.0) | (91.4) | (95.2) | (91.2) | (90.9) | (90.4) | (91.4) | (100.0) |
| **CT (n)** | 52 | 13 | 24 | 11 | 4 | 0 | 19 | 13 | 242 | 80 | 53 | 18 | 0 |
| **(%)** | (6.4) | (7.6) | (7.5) | (7.1) | (10.8) | (0.0) | (8.2) | (4.8) | (8.5) | (8.8) | (9.6) | (8.2) | (0.0) |
| **TT (n)** | 2 | 0 | 1 | 1 | 0 | 0 | 1 | 0 | 9 | 3 | 0 | 1 | 0 |
| **(%)** | (0.3) | (0.0) | (0.3) | (0.7) | (0.0) | (0.0) | (0.4) | (0.0) | (0.3) | (0.3) | (0.0) | (0.5) | (0.0) |
| **MAF** | 0.034 | 0.039 | 0.040 | 0.042 | 0.054 | 0.000 | 0.045 | 0.024 | 0.046 | 0.047 | 0.048 | 0.045 | 0.000 |
| **HWE *P*** | 0.244 | 1.000 | 0.409 | 0.231 | 1.000 | 1.000 | 0.377 | 1.00 | 0.191 | 0.448 | 0.629 | 0.364 | 1.000 |

Minor allele frequency (MAF), Hardy Weinberg Equilibrium (HWE), New Zealand (NZ), Cook Islands (CI).

The mean concentration and distributions of HDL-C and LDL-C (mmol/L) were not different between the Pacific nation subgroups or the Ngāti Porou Hauora group in the Aotearoa NZ cohort (Table 1 and Supplementary Figure 1 (post-hoc Tukey test p > 0.05, all datasets)). To test if rs1597000001 associates with HDL-C and LDL-C levels, we carried out a linear model for association of rs1597000001 adjusted by age, sex, principal component vectors (PCs) and relatedness in the Aotearoa NZ dataset (Figure 2). Significantly higher HDL-C levels were evident in those with the rs1597000001 T allele, and this association was observed in all of the surveyed Pacific nation subgroups (Aotearoa NZ NZ Māori (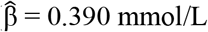 [95% CI 0.291; 0.488]), Aotearoa NZ Cook Island Māori (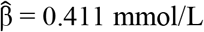 [95% CI 0.200; 0.622]), Aotearoa NZ Samoan (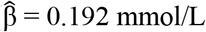 [95% CI 0.065; 0.319]), Aotearoa NZ Tongan (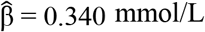 [95% CI 0.154; 0.526]), and Aotearoa NZ Other/Mixed Pacific (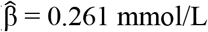 [95% CI 0.108; 0.414])) with the exception of the Aotearoa NZ Niuēan subgroup where there was no evidence of association (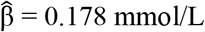 [95% CI -0.138; 0.494]) (Figure 2A). An association with HDL was also observed in the Ngāti Porou Hauora group (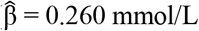 [95% CI 0.100; 0.421]).

**Figure 2.**
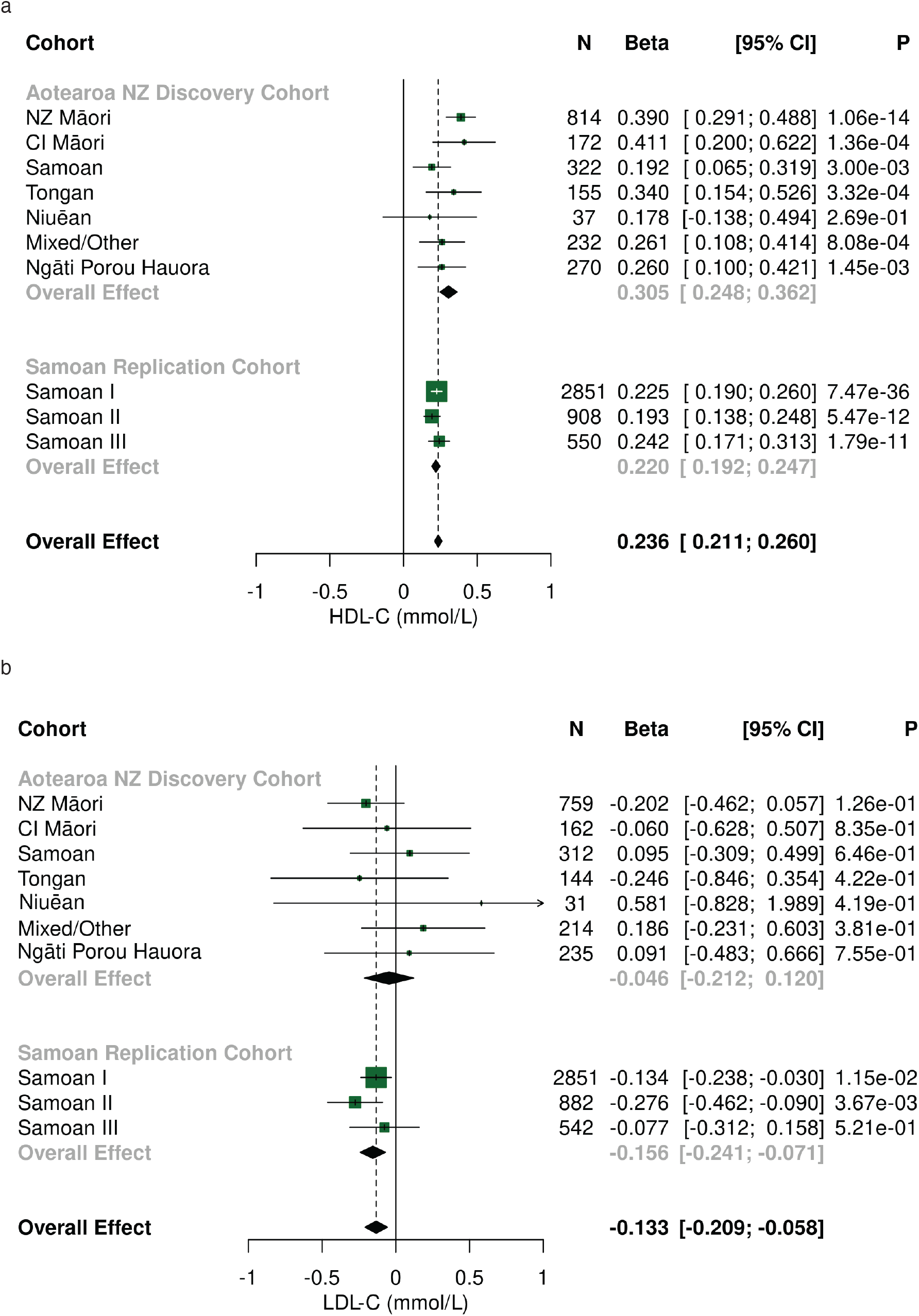
Association analyses of rs1597000001 with HDL-C and LDL-C levels in Aotearoa NZ discovery cohort and the Samoan I, Samoan II and Samoan II replication cohorts. Forest plot of a fixed-effect meta-analysis for the association of rs1597000001 with HDL-C (mmol/L) (**A**) and LDL-C (mmol/L) (**B**). Associations were adjusted by age, sex, 10 PCs and relatedness in the Aotearoa NZ cohort and age, sex, first four PCs and relatedness in the Samoan I-III cohorts. HDL-C, high-density lipoprotein cholesterol. LDL-C, low-density lipoprotein cholesterol. NZ Māori, New Zealand Māori. CI Māori, Cook Island Māori. PCs, principal components.

A fixed-effect meta-analysis showed a significant overall association of rs1597000001 T-allele with higher HDL-C (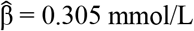 [95% CI 0.2489; 0.362]), with no evidence of heterogeneity between the sample sets (*P* = 0.223) (Figure 2A). The proportion of variance in HDL-C levels explained by rs1597000001 in the Aotearoa NZ cohort was 4.5%, similar to the proportion of variance explained by sex (5.0%). There was no association of rs1597000001 with LDL-C (Figure 2B) in any of the Pacific nation subgroups, Ngāti Porou Hauora group or in a fixed effect meta-analysis of the Aotearoa NZ cohort (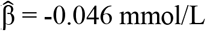 [95% CI -0.212; 0.120] (Figure 2B)).

We carried out association analyses in three independent Samoan cohorts (Samoan I-III) which serve as replication cohorts to validate the associations we observed in the Aotearoa NZ discovery cohort. Association analyses adjusted by age, sex, PCs and relatedness confirmed the association between HDL-C and rs1597000001 (Samoan cohort I: 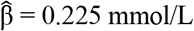 [95% CI 0.190; 0.260], Samoan cohort II: 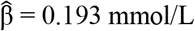 [95% CI 0.138; 0.248] and Samoan cohort III: 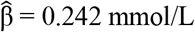 [95% CI 0.171; 0.313] (Figure 2A)) with no evidence of heterogeneity between the sample sets (*P* = 0.502). Unlike the Aotearoa NZ cohort there was an association of rs1597000001 with lower LDL-C in Samoan cohorts I and II (Samoan cohort I: 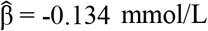 [95% CI -0.238; -0.030] and Samoan cohort II: 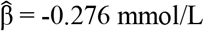 [95% CI -0.462; - 0.090] (Figure 2B)) but not Samoan III (Samoan cohort III: 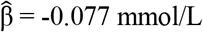 [95% CI -0.312; 0.158] (Figure 2B)). The MAF was 4.4% in Samoan I, 4.7% in Samoan II and 4.8% in Samoan III (compared to 4.0% in the Aotearoa NZ Samoan cohort) and did not deviate from HWE (*P* > 0.05, Samoan I-III (Table 2)). The proportion of variance of HDL-C explained by rs1597000001 in the Samoan I and Samoan II and Samoan III cohorts was 5.1%, 6.4% and 7.8% respectively.

A fixed-effect meta-analysis of the Aotearoa NZ Pacific nation subgroups, Ngāti Porou Hauora group and Samoan I-III cohorts demonstrated the strong association of rs1597000001 with HDL-C levels (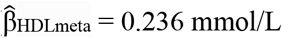 [95% CI 0.211; 0.260]) with no heterogeneity between the cohorts (*P* = 0.054). rs1597000001 associated with lower LDL-C levels in the fixed-effect meta-analysis of the Aotearoa NZ and Samoan cohorts I-III (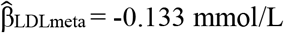 [95% CI - 0.209; -0.058]) (Figure 2B) with no heterogeneity between the cohorts (*P* = 0.534).

Next we explored what effect this variant has on HDL-C and LDL-C levels in a cohort of young Pacific people (PTO cohort) before onset of metabolic disease indicators (Table 1). The mean concentration of HDL-C was significantly higher in the PTO cohort compared to all Pacific nation subgroups and the Ngāti Porou Hauora group in the Aotearoa NZ cohort (Table 1 and Supplementary Figure 1A (post-hoc Tukey test *P* = < 0.05; all datasets)). LDL-C (mmol/L) was significantly lower in the PTO cohort in comparison to all Pacific nation subgroups and the Ngāti Porou Hauora group in the Aotearoa NZ cohort (Table 1 and Supplementary Figure 1B (post-hoc Tukey test *P* = < 0.05; all datasets)). rs1597000001 was monomorphic for the major C allele in Pacific participants without Polynesian ethnicity (i.e. of Melanesian and/or Micronesian ethnicity) (Table 2). In PTO participants with Polynesian ethnicity the MAF was 4.5% and did not deviate from HWE (*P* > 0.05). Association analysis (adjusted by age, sex, and grandparental ethnicity) in participants of Polynesian ethnicity indicated that rs1597000001 associates with higher HDL-C levels (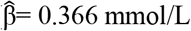 [95% CI 0.221; 0.511] (Figure 3A)) and lower LDL-C levels (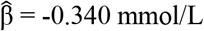 [95% CI -0.663; -0.016] (Figure 3B)) consistent with the negative estimates from the Samoan I and Samoan II cohorts. The proportion of variance of HDL-C explained by rs1597000001 in the PTO cohort was 11.8%.

**Figure 3.**
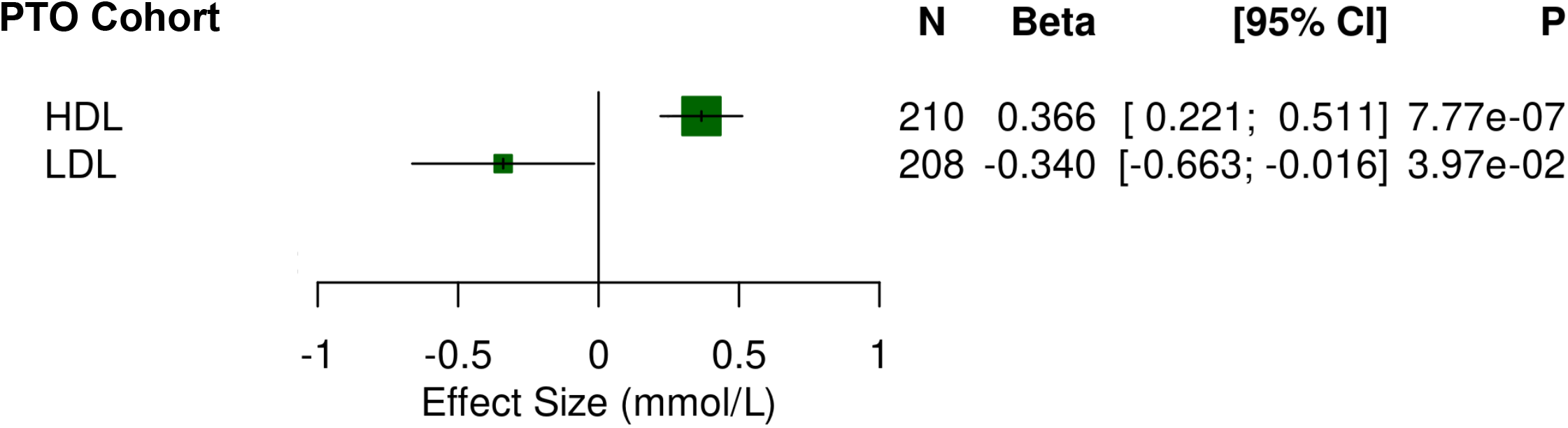
Association analyses of rs1597000001 with HDL-C and LDL-C levels in Polynesian participants of the PTO cohort. Forest plot of rs1597000001 association with HDL-C (mmol/L) and LDL-C (mmol/L) in the PTO cohort (Polynesian ethnicity only). Associations were adjusted by age, sex and grandparental ethnicity. HDL-C, high-density lipoprotein cholesterol. LDL-C, low-density lipoprotein cholesterol.

### rs1597000001 associates with decreased CETP activity

To test the hypothesis that rs1597000001 alters CETP function, we carried out CETP activity assays in serum from participants of the Pacific Trust Otago cohort who were heterozygous for rs1597000001 (n = 4) and homozygous for rs1597000001 C allele (n = 7). The rs1597000001 T-allele was significantly associated with lower activity of CETP in comparison to the homozygous carriers of the major C allele (unpaired t-test with Welch’s correction p = 0.028) (Figure 4).

**Figure 4.**
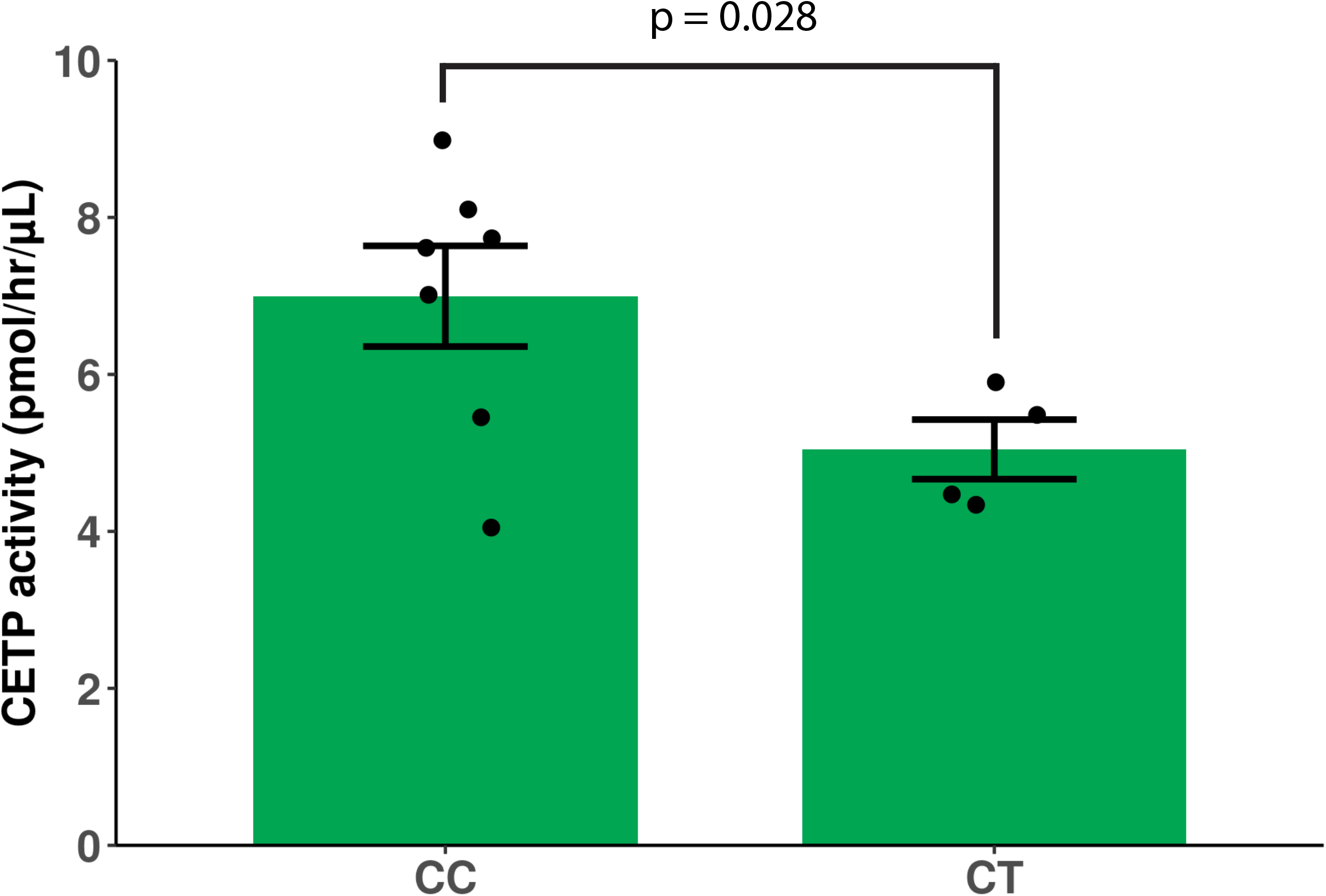
CETP activity by rs1597000001 genotype. Plasma activity of CETP (pmol/hr/μL) in 11 participants of the PTO cohort who had the heterozygous rs1597000001 genotype (CT) versus the homozygous rs1597000001 genotype for the major allele (CC). CETP, cholesteryl ester transfer protein. Error bars represent standard error of the mean. A two-sample t-test with Welch’s correction was conducted to test for a statistically significant (*P* = 0.028) difference in mean CETP activity between the two genotypic groups.

### Multiple independent genetic associations are identified at CETP in Māori and Pacific People

Genetic contribution to HDL-C is polygenic, and patients with extreme HDL concentrations possess both rare large-effect variants and common small-effect variants ^28^ at *CETP* that contribute to HDL-C ^29^. We re-examined ^15^ the entire *CETP* locus for association with HDL-C levels in Māori and Pacific peoples in the Aotearoa NZ cohort. rs1597000001 was the maximally associated variant followed by the *CETP* promoter polymorphism ‘-629 C/A’ rs1800775 (Figure 5A). The rs1800775 C-allele was present in the Aotearoa NZ cohort (46.0%) at a frequency similar to Europeans (48.0% in gnomAD), and exhibited weak linkage disequilibrium with rs1597000001 (r^2^ = 0.032). The effect size for rs1800775 C-allele (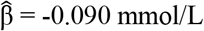 [95% CI 0.112; -0.069] was small in comparison to the effect of rs1597000001 (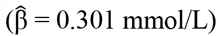 mmol/L) and the proportion of variance of HDL-C explained by rs1800775 in the Aotearoa NZ cohort was 3.3% compared to 4.5% for rs1597000001. The rs1800775 C-allele associates with the same direction of effect previously observed for Europeans (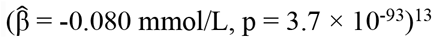, p = 3.7 × 10^−93^)^13^ and Pacific peoples (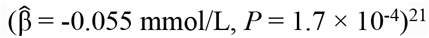, *P* = 1.7 × 10^−4^)^21^.

**Figure 5.**
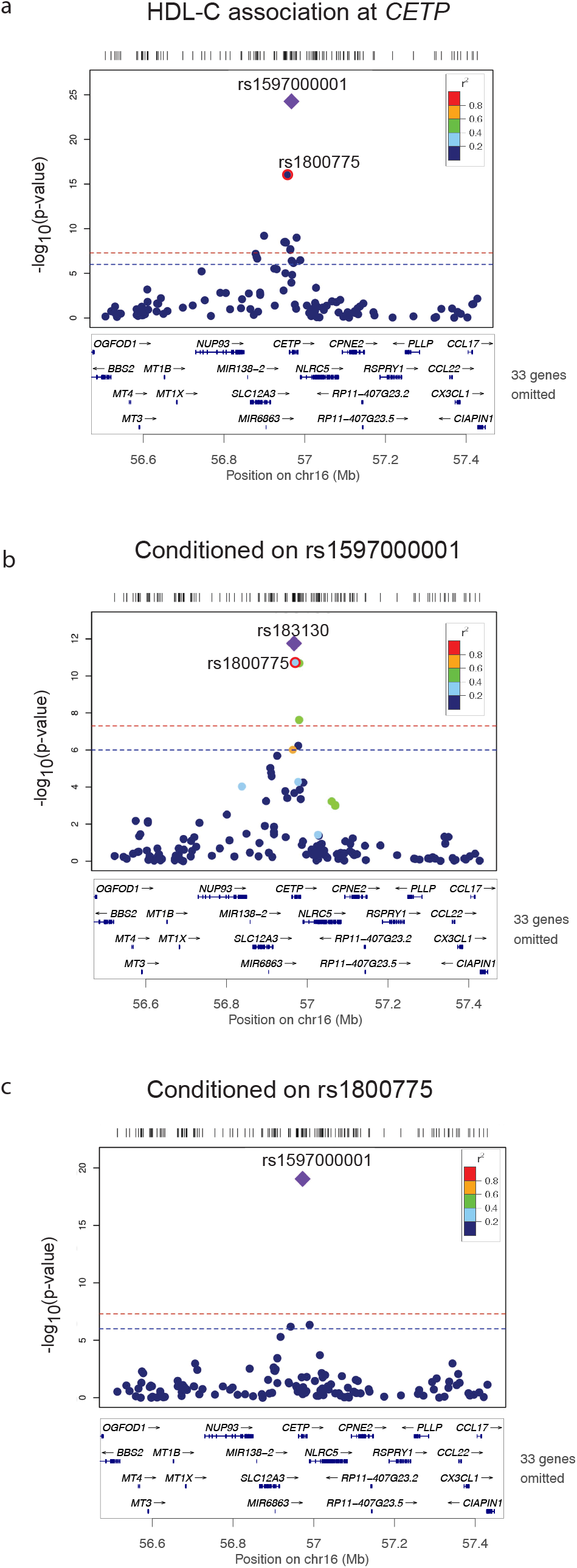
Association of HDL-C levels at the *CETP* locus. Association of HDL-C at the *CETP* locus (+/-500kb the lead variant rs1597000001) (A) and after conditioning on rs1597000001 (B) and rs1800775 (C) using variants on Illumina Infinium CoreExome v24 bead chip platform for genotyped participants from the Aotearoa NZ cohort. The strength of LD, as measured by the r^2^, between each variant and rs1597000001 (A and C) and rs183130 (B) is represented by the color of each point according to the legend in the top right hand corner. The plot was generated using a custom locus zoom R package (https://github.com/Geeketics/LocusZooms). CETP, cholesteryl ester transfer protein. HDL-C, high-density lipoprotein cholesterol.

Given the large effect of rs1597000001 on HDL-C and the fact that rs1597000001 and rs1800775 exhibit some linkage disequilibrium we carried out conditional analyses to test whether the effects at rs1800775 and rs1597000001 were dependent on each other. In the Aotearoa NZ cohort, the effect on HDL-C persisted for rs1597000001 (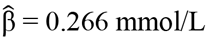 [95% CI 0.210; 0.323]) conditioned on rs1800775 genotype (Figure 5B) and for rs1800775 conditioned on rs1597000001 genotype (rs1800775 C-allele; 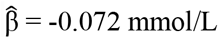 [95% CI - 0.094; -0.052]) (Figure 5C). However, after conditioning on rs1597000001, rs183130 was the most significantly associated variant (T-allele; 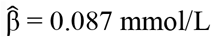 [95% CI 0.062; 0.112]) at *CETP* (r^2^ with rs1800775 = 0.31). In the Samoan I cohort, conditioning on rs1597000001 resulted in two additional signals marked by rs11076175 (G-allele; 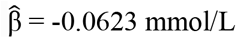 [95% CI -0.0876; -0.0369]) and rs4783961 (A-allele; 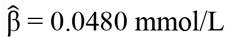 [95% CI 0.0279; 0.0681]) (Supplementary Figure 2). rs4783961 exhibited linkage disequilibrium with rs1800775 (r^2^ = 0.227) and rs183130 (r^2^ = 0.623). However, rs11076175 had only weak linkage disequilibrium with rs1800775 (r^2^ = 0.114) and rs183130 (r^2^ = 0.014).

## DISCUSSION

Using whole-genome sequence data from individuals of Māori and Pacific ethnicity, we have identified a Population-specific missense variant in *CETP* that strongly associates with a favorable lipid profile (higher HDL-C and lower LDL-C) in a meta-analysis of four independent Pacific cohorts. The positive effect on HDL-C levels of the rs1597000001 T-allele was observed in every group analysed, with the exception of the Niuēan Pacific nation subgroup. This is likely a reflection of the small sample size of this subgroup, limiting our detection power. However, the effect size estimate was positive, which is consistent with all the other island nation subgroups in the Aotearoa NZ cohort. Given the likely, but unstudied, nutritional and environmental variations across these groups within Aotearoa NZ and between Aotearoa NZ and the Samoan Islands, it is noteworthy that similar effect sizes were detected.

The effects of rs1597000001 on LDL-C are less conclusive. In the Samoan I and II cohorts, an association with lower LDL-C was observed. However, the Aotearoa NZ, Samoan III, and PTO cohorts did not show an association of rs1597000001 with LDL-C levels. The differences could be due to unmeasured heterogeneity in medication use and other genetic and environmental factors. Our findings that rs1597000001 associates with HDL-C consistently throughout Pacific nation groups but not LDL-C corroborate data from Mendelian randomization studies of CETP which indicate that CETP activity is an important causal determinant of HDL-C^30^ levels but not LDL-C levels.

Variants in *CETP* that cause loss-of-function and consequently have large effects on HDL-C levels are rare in the general population and are enriched in study populations that have high HDL-C levels ^31-33^. Conversely rs1597000001 (MAF ∼ 2.4%-5.4%) is a common variant of large effect in Māori and Pacific peoples, and the allele frequency was stable throughout the Pacific nations surveyed here. It is notable that the variant was not detected in people of Pukapukan ethnicity and Pacific peoples without Polynesian ethnicity. The most probable explanation is that the absence of the variant from people of Pukapukan ethnicity is the result of a different population history for this group for example bottle-necking event(s) and/or founder effects. That we did not observe the variant in non-Polynesian Pacific people suggests that this is a Polynesian-specific variant.

The large effect on HDL-C level that we observe for rs1597000001 is reminiscent of that observed for the common hyperalphalipoproteinemia 1 (HALP1) (elevated HDL-C levels) loss-of-function variant rs2303790 (p.Asp259Gly; 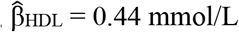)^20^ located in exon 15 of *CETP*. Functionally rs2303790 reduces CETP activity, likely emulating Anacetrapib inhibition of CETP ^34^. Anacetrapib on average raises HDL-C levels by 1.12 mmol/L in patients with atherosclerotic vascular disease ^35^. Given that rs1597000001 is located in exon 6, it is likely that its functional effect on CETP is disparate to rs2303790. However, it is notable that the effect of one allele of rs1597000001 on HDL-C is ∼25% of that observed for Anacetrapib. Here, we showed that heterozygous carriers of rs1597000001 have lower CETP activity which supports our hypothesis that the rs1597000001 T-allele results in dysfunctional CETP and consequently increases HDL-C.

The magnitude of effect on HDL-C of rs1597000001 is also comparable to carrier status for protein truncating variants in *CETP* (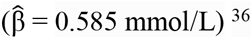) ^36^. We note that rs1597000001 is in very close proximity to a rare truncating variant p.Trp179* which introduces a stop codon in exon 6. This rare variant has only been identified in people with HALP1 ^31^ thus we are unable to make comparisons of the estimates of effect on HDL. p.Trp179* could possibly alter CETP levels through nonsense-mediated decay of the truncated mRNA. Alternatively, p.Trp179* could also functionally disrupt the CETP protein. p.Trp179* and rs1597000001 (p.Pro177Leu) are located in the N-terminal β-barrel domain of CETP which is predicted to penetrate the HDL surface ^37^, and is a putative entry site for cholesterol ester. Both variants are contained in the Ω6 flap (Gln^155^ – Trp^162^) which separates from the Ω5 flap upon HDL penetration, then opens the N-terminal distal end for uptake of cholesterol ester ^37^. Specifically, Trp^162^ plays an important anchoring role in the penetration and settling of CETP in HDL. Binding of CETP to the HDL substrate occurs via hydrophobic interactions ^38^, and it could therefore be predicted that the p.Pro177Leu substitution from proline to the hydrophobic amino acid leucine at this site could alter substrate binding and therefore cholesterol ester transfer activity of CETP. Alternatively, proline residues commonly form bends and turns in protein structure ^39^, thus the substitution to leucine could disrupt the conformation of the CETP N-terminal domain.

Although dyslipidaemia is prevalent in people of Māori and Pacific ethnicity, our study has identified a population-specific variant that associates with an improved lipid profile. It remains to be seen whether this variant is also associated with lower cardiovascular event risk. Although the variant has a tangible biological mechanism of effect and results in CETP deficiency, understanding how this variant is modified by other common, smaller-effect variants within the locus and how these variants function will be important information for more effective targeted interventions with pharmaceuticals ^40^. Most importantly, this variant and the accumulating evidence of the presence of other Population-specific trait-associated variants ^41-45^ emphasizes the significance of population-specific variation and its influence on disease traits and risk. Understanding the genetic basis of metabolic disease in Māori and Pacific populations is essential for the improvement of health outcomes for these groups ^10^, and comprehensive evaluations of genome-wide genetic variation is long overdue and critical for equity in healthcare as we move towards an era of personalized medicine grounded in genetics.

## METHODS

### Study cohorts

Demographic and biochemical characteristics of the participants are summarized in Table 1. 2,272 participants of Māori and Pacific ethnicity were recruited in Aotearoa New Zealand (NZ). The Aotearoa NZ cohort serves as the discovery cohort in this study. 4,309 participants of Samoan ethnicity were recruited in the Independent State of Samoa and the US territory of American Samoa into the Samoan I (n = 2,851), II (n = 908) and III (n = 550) cohorts which serve as the replication cohorts in this study. Finally, an additional 255 young people (aged 14 – 25 years) identifying with at least one Pacific Island ethnicity within Oceania (i.e., Melanesia, Micronesia or Polynesia) residing in Dunedin, New Zealand were recruited by the Pacific Trust Otago into the Pacific Trust Otago (PTO) Cohort.

For all participants information obtained at recruitment included age, sex, height, and weight, as measured by trained assessors. BMI was calculated by dividing an individual’s weight by the square of their height in meters. Blood biochemical measurements including lipid measurements were performed at Southern Community Laboratories (Dunedin, NZ) for the Aotearoa NZ participants, at Northwest Lipid Labs (Seattle, WA, USA) for the Samoan I cohort, and at Lipids Research Clinic at Miriam Hospital, Brown University for the Samoan II and III cohorts.

The Aotearoa NZ cohort is the amalgamation of three separate groups. 2002 participants aged ≥ 16 years, located primarily in Auckland and Christchurch were recruited to Genetics of Gout, Diabetes and Kidney Disease in Aotearoa NZ Study^41^. The participants from this study were separated into subgroups based on the self-reported Pacific nation of ethnicity of their grandparents. Those participants who also reported some non-Pacific grandparent ethnicity were grouped according to their Pacific nation ethnicity. This resulted in six sample sets: Aotearoa NZ NZ Māori (*n* = 814), Aotearoa NZ Cook Island Māori (*n* = 172), Aotearoa NZ Samoan (*n =* 322), Aotearoa NZ Tongan (*n =* 155), Aotearoa NZ Niuēan (*n =* 37) and an ‘Other/Mixed’ Aotearoa NZ Pacific group (*n =* 232), which included individuals of Tahitian (*n =* 1) and Tuvaluan (*n* = 5) ethnicity, along with individuals who self-reported grandparental ethnicity from more than one Pacific nation (*n =* 230). An additional 270 participants from Te Tairāwhiti (east coast of the North Island, NZ) were recruited in collaboration with the Ngāti Porou Hauora (Health Service) Charitable Trust. At the request of Ngāti Porou Hauora (NPH) these participants were analyzed separately to the participants in the six Pacific nation subgroups. 72 participants of Pukapukan ethnicity were recruited in collaboration with the Pukapukan Community of New Zealand Inc. in Mangere, South Auckland, NZ.

The Samoan I cohort consists of 2,851 Samoan adults aged 22–65 years residing in the Independent State of Samoa. The Samoan II cohort is a family study consisting of 908 Samoan adults aged 18-88 years residing in the Independent State of Samoa and the US territory of American Samoa. The Samoan III cohort consists of 550 Samoan adults aged 29–88 years residing in the Independent State of Samoa and the US territory of American Samoa. With the exception of the participants in the Samoan II cohort, participants were also asked about the ethnicity of each of their grandparents. Participants from the Samoan I and III cohorts all reported four Samoan grandparents. Although participants from the Samoan II cohort did not report the ethnicity of their grandparents, in principal components analysis of ancestry (see below), the participants of this cohort cluster together with participants from the Samoan III cohort.

Ethical approval for the Aotearoa NZ cohort study was given by the NZ Multi-Region Ethics Committee (MEC/05/10/130; MEC/10/09/092; MEC/11/04/036), the Northern Y Region Health Research Ethics Committee (Ngāti Porou Hauora Charitable Trust study; NTY07/07/074), and the University of Otago Human Health Ethics Committee (Pacific Trust Otago; 12/349). Ethical approval for the Samoan I cohort study was given by the Health Research Committee of the Samoa Ministry of Health and the institutional review board of Brown University. Ethical approvals for the Samoan II and III cohort studies were given by the Health Research Committee of the Samoa Ministry of Health and the Institutional Review Boards (IRBs) of the Department of Health in American Samoan; of The Miriam Hospital, Providence, Rhode Island; and of Brown University. The consent forms for Samoan I, II, and III cohorts were available to participants in both Samoan and English. All participants provided written informed consent for the collection of samples and subsequent analysis. Participants recruited in Aotearoa New Zealand were asked if they would like a karakia (Māori prayer) carried out upon disposal of their blood samples, if indicated this was carried out. The PTO project was guided by the University of Otago Pacific Research Protocol.

### Identification of rs1597000001

For genomic sequencing 2 μg of total genomic DNA from 55 Māori and Pacific individuals (participants with gout in Ardea Biosciences (USA) clinical trials for urate-lowering therapy ^42,46,47^) were submitted to Kinghorn Centre for Clinical Genomics at Garvan Institute of Medical Research in New South Wales, Australia for library preparation and Next Generation Sequencing 30x WGS (TruSeq Nano v2.5). The *CETP* gene region (based on reference transcripts from Ensembl (https://asia.ensembl.org/index.html)) was extracted from whole genome sequence data in FASTQ format aligned to the human genome (GRCh37/hg19) following implementation of the Genome Analysis ToolKit (GATK) best practices using the Burrows-Wheeler Aligner ^48^Picardtools (https://broadinstitute.github.io/picard/) and GATK v3.6.0 ^49^ (NeSI pipeline for GATK; DOI: http://doi.org/10.5281/zenodo.2564243). Variants in *CETP* were annotated with allele frequencies from the Genome Aggregation Database (http://gnomad.broadinstitute.org/) ^23^ to identify population-specific variants, and annotated using Variant Effect Predictor (VEP) (www.ensembl.org/Tools/VEP) ^50^. *In silico* predictions for deleteriousness of rs1597000001 and conservation at rs1597000001 were obtained using the UCSC browser (GRCh37/hg19) track collection ^51^ for Combined Annotation Dependent Depletion (CADD) scores ^24^ and Genomic Evolutionary Rate Profiling (GERP) scores for Mammalian Alignments ^25^ respectively. The CETP protein structure (10.2210/pdb2OBD/pdb) ^26^ was obtained from The Protein Databank (https://www.rcsb.org/) ^52^ and visualized using the PyMOL molecular graphics system (v2.3.2 (Shrodinger, LLC)).

### Genotyping

In the Aotearoa NZ, NPH and PTO cohorts, rs1597000001 was genotyped using a custom-designed TaqMan probe-set (Applied Biosystems, Foster City, CA, USA). A custom Python script (snp_design; DOI: https://doi.org/10.5281/zenodo.56250) was used to annotate the human genome build 37 reference sequence (ftp://ftp.ensembl.org/pub/grch37) with rs1597000001 and any surrounding SNPs (obtained from the NCBI dbSNP build 147 common SNP list; ftp://ftp.ncbi.nlm.nih.gov/snp) before primer and probe design. Forward primer: CGGTGCCTGGTACACACTAG; reverse primer: TGTGAACAGCTGCTTGATCCA; probe 1 (VIC): CTGCCTTCAGGCCTG; probe 2 (FAM): CTGCCTTCAGGCTTG. Genotyping was carried out using the LightCycler 480 Real-Time Polymerase Chain Reaction System (Roche Applied Science, Indianapolis, IN, USA) in 384 well plates. Additional genotypes beyond rs159700001 at the *CETP* locus for the Aotearoa NZ participants were obtained from the Illumina Infinium CoreExome v24 bead chip platform whole-genome genotyping data generated as previously described ^41^. In the Samoan I cohort, participants were genotyped using the Genome-Wide Human SNP Array 6.0 (Affymetrix, CA, USA) with quality control as described in ^53^. rs1597000001 was not included on the array, but the genotypes from the array served as the scaffold for imputation of the variant. A subset of Samoan I cohort participants (*n* = 1,294) were whole-genome sequenced as part of the Trans-Omics for Precision Medicine (TOPMed) program ^54^, a Samoan-specific haplotype reference panel was generated from the sequences using Eagle2 (v.2.3.5) ^55^, and additional genotypes, including those of rs1597000001 were imputed based on the genotype scaffold and the reference panel using Minimac4 ^56^. In the Samoan II and III cohorts, participants were genotyped using the Infinium Global Screening Array-24 v3.0 BeadChip (Illumina, CA, USA) with custom content that included rs1597000001. Quality control and quality assurance checks for the Samoan II and III cohorts were conducted using GWASTools ^57^.

### Principal components and relatedness matrices

Whole-genome principal component vectors were calculated for the Aotearoa NZ cohort using 2,858 ancestry informative markers (as identified by Illumina) extracted from the CoreExome whole-genome genotypes. Ten PCs were generated from the SmartPCA (EIGENSOFT v6.0.1)) ^58^ program and used as covariates in the association analyses to account for population stratification and cryptic relatedness. Relatedness coefficients were calculated in the Aotearoa NZ dataset using the software GEMMA (v0.98.4) ^59^ from 257,069 independent SNPs from the CoreExome whole-genome genotyping data.

For the Samoan I, II, and III cohorts, principal components and empirical kinship coefficients were calculated using genotypes from the Genome-Wide Human SNP 6.0 array (Samoan I) and the Infinium Global Screening Array-24 v3.0 BeadChip (Samoan II and III). In the Samoan I cohort, PCs were calculated as described in Minster *et al*. ^53^ and the empirical kinship matrix was calculated from 10,000 independent autosomal markers using OpenMendel ^53,60,61^. In the Samoan II and III cohorts, 55,640 autosomal markers were used to calculate PCs and empirical kinship matrices with PC-AiR and PC-Relate, respectively ^62^, per the recommended procedure in the GENESIS R/Bioconductor package ^63^.

### Statistical analyses

All association analyses described below were carried out using the R v4.0.2 software. For the association analyses in the Aotearoa NZ cohort, a generalized linear mixed model-based Wald test was carried out using GMMAT software (v1.3.1) ^64^ to test for associations between rs1597000001 and the continuous variables HDL-C and LDL-C. Analyses were adjusted by sex, age, 10 PCs and relatedness. A linear model was used to test for associations in the PTO cohort between rs1597000001 and the continuous variables HDL-C and LDL-C using the `lm()` function in R. The PTO analyses were adjusted by sex, age and self-reported ethnicity group for each grandparent, owing to absence of whole-genome ancestry informative markers in this cohort. For the association analyses in the Samoan I cohort, linear mixed-model regression of the phenotypes on rs1597000001 was performed using the `lmekin()` function of the coxme R package with sex, age, four PCs and relatedness as covariates. In the Samoan II and III cohorts, a mixed model association testing was carried out using `lmekin()` to test for associations between rs1597000001 and HDL-C or LDL-C phenotypes with sex, age, first four PCs, polity (Samoa or American Samoa), and relatedness as adjusting variables.

Each Pacific population sample set (Aotearoa NZ cohort: Aotearoa NZ NZ Māori, Aotearoa NZ Cook Island Māori, Aotearoa NZ Samoan, Aotearoa NZ Niuēan, Aotearoa NZ Pukapukan and Aotearoa NZ ‘Other /Mixed’ Pacific nations subgroups; Ngāti Porou Hauora group, Samoan Cohort I, Samoan Cohort II and Samoan Cohort III; and Pacific Trust Otago) was analyzed separately, and the effects combined for the Aotearoa NZ cohort and all cohorts (excluding the PTO cohort) in an inverse variance-weighted fixed-effect meta-analysis using the R package meta (v.3.0-2). Heterogeneity between sample sets was assessed using Cochran’s heterogeneity (Q) statistic. The proportion of variance explained by rs1597000001 and *CETP* promoter common variant rs1800775 for HDL-C was calculated using the `partial.rsq()` function in the rsq R package (v2.2) ^65^ which calculates the partial R-squared value for each predictor from the linear model generated by the `lm()` function in R. The β coefficient in all analyses represents the estimated effect on HDL-C and LDL-C units (mmol/L) per copy of the rs1597000001 T-allele. For the Aotearoa NZ analyses HDL-C and LDL-C measurements were in mmol/L. In the Samoan cohorts I, II and III measurements were in mg/dl and converted to mmol/L before the association analyses.

To contextualize the effect of rs1597000001 among the other variation in *CETP*, variants on the Illumina Infinium CoreExome v24 bead chip platform and present in the Aotearoa NZ cohort (minor allele frequency (MAF) > 0.01) were extracted from the *CETP* region (rs1597000001 ± 500 kb). A linear model was used to test variants for association with HDL-C using PLINK (v1.90b6.10) adjusted by age, sex and 10 PCs. Linkage disequilibrium and conditional analyses for rs1597000001 and *CETP* promoter common polymorphism rs1800775 in the Aotearoa NZ cohort were carried out in PLINK (v1.90b6.10), adjusting by age, sex and 10 PCs calculated from the Aotearoa NZ dataset. Conditional analysis was conducted in Samoan cohort I across the *CETP* region with rs1597000001 modeled as an additional fixed-effect covariate. Regional association plots were created using a custom R package (LocusZoom-like Plots: DOI: https://doi.org/10.5281/zenodo.5154379) for the Aotearoa NZ cohort and using LocusZoom ^66^ for the Samoan I cohort.

### CETP activity

Individuals of the PTO cohort were recalled specifically for the purpose of providing fresh plasma samples for the CETP activity assays. CETP activity in 11 participants, of whom four were heterozygous for rs1597000001 and seven were homozygous for rs1597000001 C-allele was assessed using the CETP activity assay kit II (F) from BioVision (K595-100) according to the manufacturer’s instructions. Fresh plasma (1 µL) was used for the assays which were carried out in technical triplicate. Fluorescence was measured using a BMG LabTech CLARIOstar plate reader and CLARIOstar software (v.5.01 R2, Firmware 1.10). Using the `stat.desc()` function of the pastecs R package, we tested whether data within each genotypic group was normally distributed. We found that both genotypic groups had skewness/2SE and kurtosis/2SE values between -1 and 1, indicating normality of the data. The Shapiro-Wilks tests for skewness were also non-significant. A two-samples t-test with Welch’s correction was conducted to test for a difference in mean CETP activity between the two genotypic groups. One technical replicate was removed after it was identified as an outlier based on a significant Grubbs test for one outlier (*P* = 0.00065) using the `grubbs.test()` function from the outliers R package. Statistical analyses were conducted in R v4.0.2 software with all data reported as means ± standard error of the mean.

## Data Availability

The data from the Aotearoa NZ and PTO cohorts are not publicly available owing to consent restrictions but can be requested from the corresponding author under an appropriate arrangement. Samoan cohort I data are available from dbGaP (accession number: phs000914.v1.p1). Samoan II and III cohort data (recruited in 2002 to 2003 and 1990 to 1995, respectively) are not available as participants were not consented for data sharing.

https://github.com/MerrimanLab/CETP-Project

## ACKNOWLEDGMENTS

The authors sincerely thank the participants for generously donating their time and information to this study. The authors would like to thank J. Drake (Department of Rheumatology, Canterbury District Health Board, Christchurch, New Zealand), J. de Kwant, R. Laurence, C. Franklin, and M. House (all Department of Medicine, University of Auckland, Auckland, New Zealand), J. Harre Hindemarsh, N. Aupouri, R. Akuhata, and C. Ford (all of Ngāti Porou Hauora Charitable Trust, Te Puia Springs, New Zealand), G. Sexton (Counties Manukau District Health Board, Auckland, New Zealand), and B. Vincent (Waitemata District Health Board, Auckland, New Zealand) for recruitment. For the Samoan cohorts, we acknowledge the Samoan Ministry of Health, the Samoa Bureau of Statistics, and the American Samoan Department of Health for their support of this research. The work was funded by the National Institute of Health grants R01HL093093 (STM), R01HL133040 (RLM), R01AG09375 (STM), R01HL52611 (MI Kamboh), R01DK59642 (STM), and R01DK55406 (RD). Molecular data for the Trans-Omics in Precision Medicine (TOPMed) program was supported by the National Heart, Lung and Blood Institute (NHLBI). Genome sequencing for the “[NHLBI TOPMed: Genome-wide Association Study of Adiposity in Samoans](http://www.ncbi.nlm.nih.gov/projects/gap/cgi-bin/study.cgi?study_id=phs000972.v4.p1)” (phs000972.v4.p1) was performed at the Northwest Genomics Center (HHSN268201100037C) and the New York Genome Center (HHSN268201500016C). Core support including centralized genomic read mapping and genotype calling, along with variant quality metrics and filtering were provided by the TOPMed Informatics Research Center (3R01HL-117626-02S1; contract HHSN268201800002I). Core support including phenotype harmonization, data management, sample-identity QC, and general program coordination were provided by the TOPMed Data Coordinating Center (R01HL-120393; U01HL-120393; contract HHSN268201800001I).

## AUTHOR CONTRIBUTIONS

MM, RT, TJM and AP-G coordinated the sampling, genome sequencing and genome-wide genotyping for the Aotearoa NZ cohort and 55 Aotearoa NZ genome sequences. MB and MC carried out the genome sequencing data analyses in the 55 genome sequences. For the Samoan I cohort NH led the field work data collection and phenotype analyses under the supervision of STM. For the Samoan II and III cohorts, STM led the field work data collection and phenotype analyses. GS and HC performed genotyping experiments for the Samoan I cohort, and HC prepared DNA for genotyping at the Center for Inherited Disease Research for the Samoan II and III cohorts and for sequencing by the TOPMed Program for the sequenced subset of the Samoan I cohort, under the supervision of RD. JZZ and JCC performed genotype imputation for the participants in the Samoa I cohort with guidance from RLM and DEW. JM coordinated the samples and genotyping for the PTO cohort and carried out the genotyping over the Aotearoa NZ cohort. JM, MK, and ML carried out the association analyses with guidance from TRM, DEW, RLM and assistance from NS, RT, EMR, JCC and TJM. MR and BM carried out the CETP assays supervised by SM. MSR, SV, and JT facilitated fieldwork in Samoa and American Samoa. TN, MSR, SV, NH, PW, FK, MT, ND, LS, RM and JdZ contributed to the discussion of the public health and cultural implications of the findings. ML wrote the manuscript with guidance from TRM and contribution from all co-authors. All co-authors contributed to this work, discussed the results, and critically reviewed and revised the manuscript.

## DATA AVAILABILITY

The data from the Aotearoa NZ and PTO cohorts are not publicly available owing to consent restrictions but can be requested from the corresponding author under an appropriate arrangement. Samoan cohort I data are available from dbGaP (accession number: phs000914.v1.p1). Samoan II and III cohort data (recruited in 2002–2003 and 1990–1995, respectively) are not available as participants were not consented for data sharing.

## CODE AVAILABILITY

All software used in the analyses were open source and described in the Methods. Code written for the analyses are available in GitHub: https://github.com/MerrimanLab/CETP-Project

## FIGURE LEGENDS

**Supplementary Figure 1.**
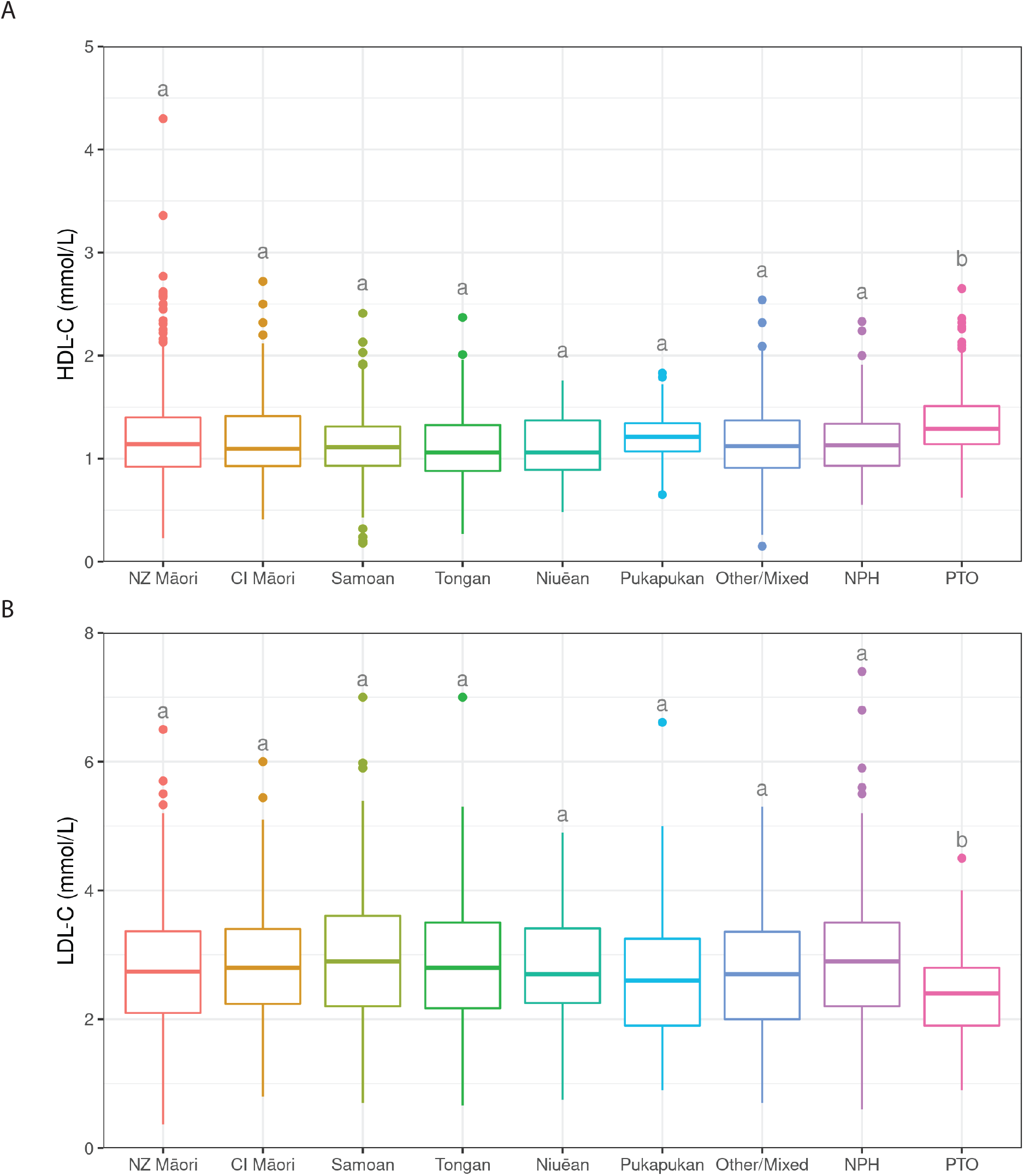
HDL- and LDL-C (mmol/L) distributions in the Aotearoa New Zealand cohort separated into Pacific nation of grand-parental ethnicity and Ngāti Porou Hauora subgroups and the Pacific Trust Otago. HDL-C, High-density lipoprotein cholesterol. LDL-C, Low-density lipoprotein cholesterol. NZ Māori, New Zealand Māori. CI Māori, Cook Island Māori. NPH, Ngāti Porou Hauora, PTO, Pacific Trust Otago. Datasets that do not share a letter are statistically significantly different (p < 0.05) as assessed by a post-hoc tukey test.

**Supplementary Figure 2.**
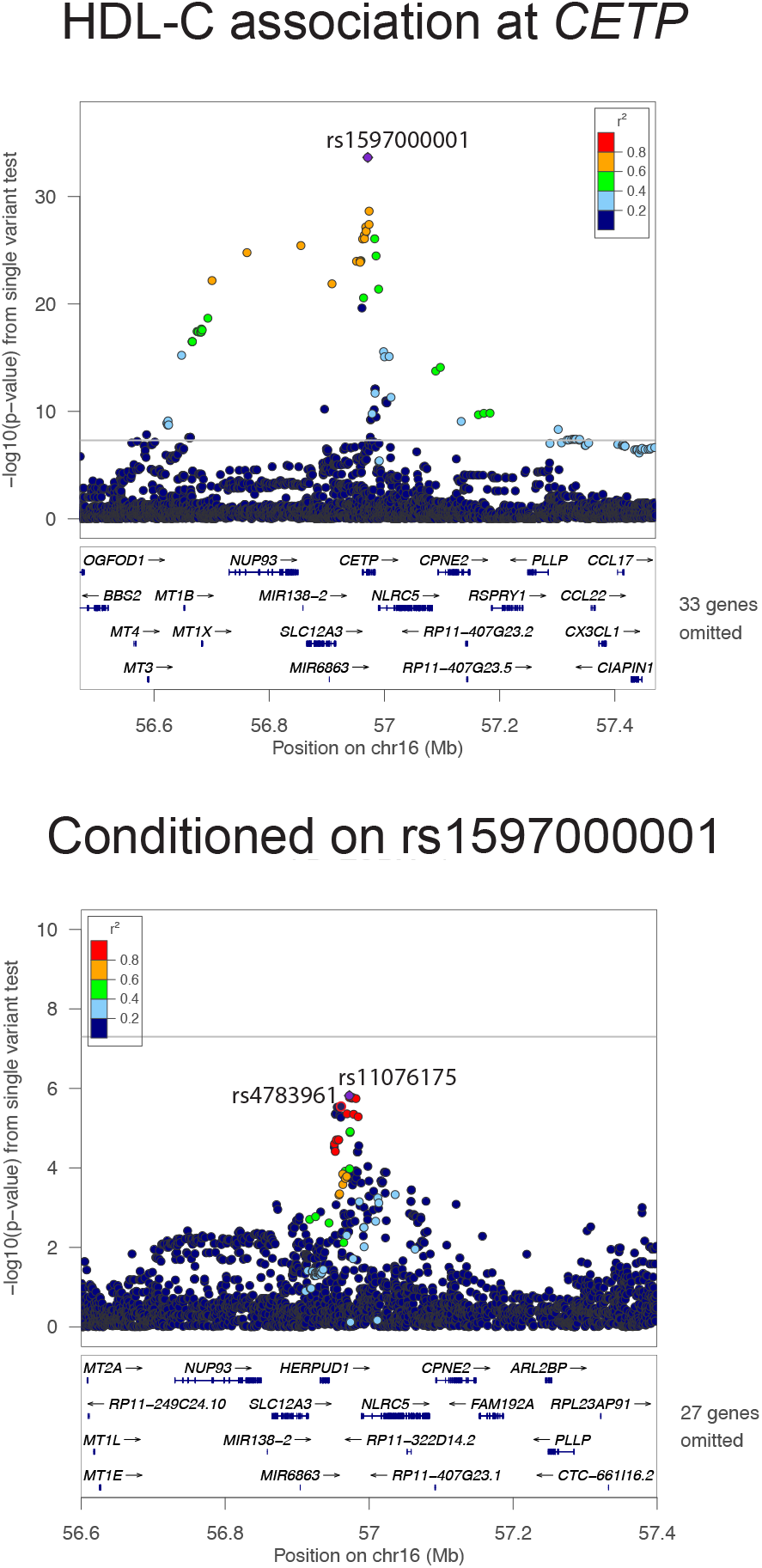
Association of HDL-C levels at the CETP locus in the Samoan I cohort. Association of HDL-C at the CETP locus (+/-500kb the lead variant rs1597000001) (A) and after conditioning on rs1597000001 (B) using using imputed data for participants from the Samoan cohort I. The strength of LD, as measured by the r2, between each variant and rs1597000001 (A) and rs11076175 (B) is represented by the color of each point according to the legend in the top right hand corner. The plot was generated using LocusZoom (Pruim et al. 2010). CETP, cholesteryl ester transfer protein. HDL-C, high-density lipoprotein cholesterol.

